# Coenrolment in a Stress Ulcer Prophylaxis Trial: Protocol for a Substudy of Characteristics and Consequences

**DOI:** 10.1101/2025.07.22.25331986

**Authors:** Diane Heels-Ansdell, Nicole Zytaruk, France Clarke, Lori Hand, William Dechert, Deborah Cook

**Affiliations:** Department of Health Research Methods, Evidence and Impact, Faculty of Health Sciences, McMaster University, Hamilton, Canada; Research Institute, St. Joseph’s Healthcare, Hamilton, Ontario; Department of Critical Care, St. Joseph’s Healthcare, Hamilton, Ontario; Department of Critical Care, Hamilton Health Sciences, General Site, Hamilton, Ontario; Department of Critical Care, Brantford General Hospital, Brantford, Ontario; Department of Medicine, Faculty of Health Sciences, McMaster University, Hamilton, Canada

**Keywords:** upper gastrointestinal bleeding, coenrolment, critically ill patients

## Abstract

**Background:** Coenrolment is defined as the enrolment of an individual patient into more than one study. This protocol describes a study of coenrolment nested in a randomized, blinded, parallel-group trial of stress ulcer prophylaxis comparing the effect of pantoprazole versus placebo on the primary efficacy outcome of clinically important upper gastrointestinal bleeding, and the primary safety outcome of 90-day all-cause mortality. The objective of this study is to determine the characteristics of patients, centers and studies involved in coenrolment, and the association of coenrolment with trial metrics and patient outcomes.

**Methods:** This is a pre-planned study with 5 specific aims and a priori hypotheses. The aims are to analyze: 1) the informed consent model of the coenrolled studies, timing of coenrolment, coenrolled study affiliation, and coenrolled study funding; 2) the characteristics of patients and centers involved in coenrolment versus not involved in coenrolment; 3) the association of coenrolment with adverse events; 4) the effect of coenrolment on protocol deviations; and 5) the impact of coenrolment on the effect of pantoprazole on the primary efficacy outcome (upper gastrointestinal bleeding) and the primary safety outcome (90-day mortality). We will use descriptive analyses and regression analysis to examine patterns and predictors of coenrolment.

**Results:** All 4821 trial participants will be included in this trial, 1719 (35.7%) of whom were coenrolled in at least one other study.

**Conclusions:** Among invasively ventilated patients in this stress ulcer prophylaxis trial, we will generate much needed empiric evidence about coenrolment in critically ill patients. These data will help to inform future guidance documents on this topic.

## Introduction

Coenrolment is the enrolment of an individual patient into more than one study [1]. For studies in the intensive care unit (ICU), coenrolment is relatively common, yet represents unique challenges [2]. Advantages of coenrolment include offering more research opportunities to participants, timely study completion, and research efficiencies. Disadvantages of coenrolment include the informed consent burden on the patient/substitute decision maker (SDM), safety risk for participants, and researcher workload. Concerns exist about mechanistic interactions when patients are enrolled in 2 interventional trials that may influence trial results or patient safety.

Building on experiences in neonatal medicine [3], resuscitation [4], pediatric critical care [5] and adult critical care [6-8], we examined this controversial issue in detail in a recent stress ulcer prophylaxis trial (REVISE [Re-Evaluating the Inhibition of Stress Erosions]) trial [9]. REVISE enrolled invasively ventilated patients ≥18 years of age to identify the effect of pantoprazole versus placebo on the primary efficacy outcome of clinically important upper gastrointestinal bleeding, and the primary safety outcome of 90-day mortality. Pantoprazole significantly reduced the risk of clinically important upper gastrointestinal bleeding and patient important upper gastrointestinal bleeding, but did not affect any other outcomes [10]. Among 4821 participants, 1719 (35.7%) were coenrolled in at least one other study.

**The overall objectives of this study were to determine the characteristics of patients, centers and studies involved in coenrolment, and the association of coenrolment with trial metrics and patient outcomes** in a randomized trial of stress ulcer prophylaxis.

## Methods

### Design

This is a pre-planned nested observational study representing a secondary analysis of the REVISE trial focused on the trial metric of coenrolment [9]. Research coordinators prospectively collected data on study-specific coenrolment variables (study name, design, affiliation, funding, enrolment timing relative to REVISE enrolment, consenting model) on trial case report forms, queried and validated at 90-days post randomization. The REVISE Methods Center retrospectively documented variables related to participating staff (e.g., critical care research experience) and center (e.g., ICU bed size).

## Participants

Patients receiving invasive mechanical ventilation who were expected to remain ventilated beyond the calendar day after randomization were eligible for REVISE. Exclusion criteria included those who had been invasively ventilated for greater than 72 hours, received acid suppression for >1 daily-dose-equivalent in the ICU, received dual antiplatelet therapy or combined antiplatelet agent and therapeutic anticoagulation, had actively or recently experienced upper gastrointestinal bleeding, or had another clear indication or contraindication to pantoprazole [9]. Eligible patients were enrolled by either a priori informed consent, deferred consent, or opt out, aligning with local ethics approvals. Patients received blinded study drug (intravenous pantoprazole 40mg daily or placebo) while invasively ventilated unless a contraindication developed for up to 90 days while in the ICU. Aligned with principles guiding non-pandemic critical care research during a pandemic [11], REVISE was paused for the shortest necessary amount of time in each participating center.

### Specific Aims

The 5 specific aims of this study will be to analyze

1. the **informed consent model** of the coenrolled studies (*a priori*, consent to continue or waived), **timing of coenrolment** (concurrent with REVISE, before or after REVISE enrolment), coenrolled **study affiliation** and coenrolled **study funding**. *We hypothesize that the dominant informed consent model would be a priori consent, that the coenrolment timing would be similarly distributed before, during and after REVISE, and that most coenrolments would involve studies affiliated with the CCCTG or ANZICS-CTG*.
2. the **characteristics of patients and centers** involved in coenrolment versus not. *We hypothesize that patients with higher illness severity and SARS-CoV-2 would be more likely to be coenrolled than other patients. We anticipate that the REVISE consent models would be similar whether patients were coenrolled or not. We hypothesize that large centers, those with more experienced research coordinators, and in Canada and Australia will be more likely to coenrol than others*.
3. the association of coenrolment with **adverse events**. *We hypothesize no association between coenrolment status and adverse events*.
4. the effect on **protocol deviations** (missed dose, wrong drug, open label proton pump inhibitor, open label histamine-2-receptor antagonist). *We hypothesize no association between coenrolment and protocol deviations*.
5. the impact of coenrolment on the effect of pantoprazole on the **primary efficacy outcome (upper gastrointestinal bleeding) and the primary safety outcome (90-day mortality)**. *We hypothesize no effect modification of coenrolment on the trial’s primary efficacy and primary safety outcomes*.

### Coenrolment Methods

Before REVISE launched and as the trial unfolded, study designs were discussed in terms of their suitability for coenrolment. In general, coenrolment was approved for any retrospective study, prospective observational study, qualitative study, mixed-methods study, survey or audit. For randomized trials, email correspondence between the REVISE Methods Center and a relevant Methods Center representative of the candidate study resulted in discussion as to whether coenrolment could impact the scientific integrity of either study. Therefore, each potential study into which a REVISE patient may be coenrolled was reviewed and reasoned collaboratively about the suitability of coenrolment. When uncertainty prevailed, the decision was made with the Steering Committee of each trial, and 2 affiliated consortia for REVISE (e.g., the Canadian Critical Care Trials Group, the Australian and New Zealand Intensive Care Society Clinical Trials Group).

Decisions were in accord with national critical care guidelines for Canada [12], Australia [13] and the UK [14], and per national or regional ethical and regulatory norms elsewhere. If coenrolment into a study was endorsed by the REVISE Methods Center, each participating center handled the opportunity for coenrolment governed by their own formal or informal coenrolment policies, institutional review board, or health authority.

Patients could have been coenrolled in another study before, concurrent with, or subsequent to enrolment in REVISE. We define sequential enrolment as enrolment of one participant into two or more studies sequentially with the consent encounters occurring in series. We define simultaneous enrolment as enrolment of one participant into two or more studies simultaneously with the consent encounters occurring concurrently.

### Statistical Analysis

Plans to examine this trial metric were outlined in the REVISE protocol [9]. The sample size for this coenrolment analysis will be fixed at 4821 participants in the REVISE Trial.

We will present data using descriptive statistics using mean and standard deviation or median and interquartile range as appropriate, or absolute counts and percentages.

To investigate the association between patient and center characteristics and coenrolment, we will perform a multilevel logistic regression analysis with coenrolment as the dependent variable. Results will be reported as odds ratios with corresponding 95% confidence intervals (CI); 2-tailed p-values will be calculated, with p< 0.05 indicating statistical significance.

To evaluate the impact of coenrolment on the effect of pantoprazole on the primary efficacy outcome (upper gastrointestinal bleeding) and the primary safety outcome (90-day mortality), we will use Cox proportional hazards regression including the following independent variables: randomized treatment, coenrolment or not, and the interaction between the two.

Regression results will be reported as hazard ratios (HR) and 95% CIs with p<0.05 indicating statistical significance.

We will report any missing data pertaining to coenrolment. We will not adjust for multiplicity. Analyses will be performed using SAS 9.4 (Cary, NC).

### Ethics and Dissemination

Protocol implementation for REVISE and database training aligned with the International Council for Harmonisation Guidelines for Good Clinical Practice and other locally applicable regulations. The REVISE trial protocol was registered on clinicaltrials.gov (NCT 03374800), was approved by Health Canada and all research ethics boards associated with participating centers.

## Results

Results of this study will generate information about the patterns, predictors and consequences of coenrolment in invasively ventilated patients randomized in a recent stress ulcer prophylaxis trial. All 4821 trial patients will be included in these analyses. In our original report, we documented that 1719 (35.7%) of patients were coenrolled in at least one other study [10].

## Discussion

This study will provide contemporary data on coenrolment in an international critical care trial conducted during the pandemic when coenrolment opportunities were highlighted [15].

The Veterans Affairs/National Institutes of Health Acute Renal Failure Trial Network found that non-pursuit of coenrolment was responsible for 2.3% of exclusions from a trial comparing renal replacement therapy strategies in critically ill patients [16]. Prohibiting coenrolment may delay trial closure, although few trials report such metrics. A cross-sectional study of trials funded between 2010-2019 in the UK National Institute for Health Research Journals Library, found that coenrolment was addressed in 94 (42.9%) of 219 publicly available protocols [17]. One quarter (23, 24.5%) did not allow coenrolment, while the rest permitted it (71, 75.5%), with a range of caveats considering the study outcomes, intervention type, and patient burden. The final decision for coenrolment rested with the local recruitment team in 57 (60.6%) and with the central organizing team in 37 (39.4%). Considering completed trials, early recruitment completion occurred in 8 of 64 (12.5%) of trials permitting coenrolment and in 5 of 20 (25.0%) where it was prohibited (p=0.285). An extension to recruitment time was required for 31 of 64 (48.4%) trials in which coenrolment was permitted and 9 of 11 (45.0%) trial for which it was not (p=0.788).

Limitations of this study include lack of information on all other studies operating in all participating centers over this 4-year trial recruitment period in relation to coenrolment. We have no information on situations in which coenrolment could have occurred but did not.

Strengths of this study include the detailed pre-specified analysis plans. The trial question was relevant for patients with and without COVID-19 [18] so both will be included in the analysis [19,20]. We will use multilevel regression to take into account clustering of patients and personnel within centers in 7 countries. This international database will enhance the generalizability of the findings.

### Knowledge Translation

We will share results with the Canadian Critical Care Trials Group and at gatherings of other national research consortia. We will publish these findings rapidly through conventional academic channels (e.g., abstracts, posters, peer-review manuscripts). We will also disseminate results at professional fora to research trainees (e.g., Masters and PhD research rounds), early career investigators (e.g., seminars), and established investigators (e.g., research consortia and conferences). We plan to work with international colleagues including those affiliated with national research consortia to generate updated global guidelines on coenrolment in the ICU setting.

## Conclusions

This study will fill an evidence void, generating information about the characteristics and consequences of coenrolment in invasively ventilated patients randomized in a recent stress ulcer prophylaxis trial.

## Data Availability

Not applicable - Protocol Manuscript

## Acknowledgements

The trial was designed by the REVISE Steering Committee including National and International Management Committees, the REVISE Investigators and Research Coordinators, the Canadian Critical Care Trials Group and the Australian and New Zealand Intensive Care Society Clinical Trials Group. We are grateful to the Methods Center teams at McMaster University (Lisa Buckingham, France Clarke, Mary Copland, Karlo Matic, Ashley Sawyer, Alyson Takaoka) and The George Institute (Fatima Butt, Anna Cheng, Conrad Nangla, Fiona Osborne, Tina Schneider, Isabella Schoeler, Anna Tippet) for their expertise.

We appreciate the work of Research Coordinators and Site Investigators on REVISE, and all the patients and family members who participated in this trial.

## Career Award Funding

Dr. D Cook holds a Research Chair in Knowledge Translation in Critical Care from the Canadian Institutes for Health Research.

## Ethics approval

REVISE was approved by Health Canada [HC6-24-c210404], Clinical Trials Ontario Research Ethics Project ID: 1360, the Northern Sydney Local Health District Human Research Ethics Committee (HREC) [2019/ETH08405], and Comissão Nacional de Ética em Pesquisa (CONEP) [5.734.590]. All participating centers had local ethics approval.

## Authors’ contributions

**Concept and design:** N Zytaruk, F Clarke, D Heels-Ansdell, D Cook

**Acquisition, analysis, or interpretation of data:** N Zytaruk, F Clarke, L Hand, W Dechert, D Heels-Ansdell, D Cook

**Drafting of the manuscript:** D Cook, D Heels-Ansdell

**Critical revision of the manuscript for important intellectual content:** N Zytaruk, F Clarke, L Hand, William Dechert

**Statistical analysis:** D Heels-Ansdell

**Obtained funding**: D Cook, N Zytaruk

**Administrative, technical, or material support:** N Zytaruk, D Heels-Ansdell, D Cook

**Data Integrity:** N Zytaruk, F Clarke, L Hand, W Dechert, D Heels-Ansdell, D Cook

**Data Sharing:** The REVISE dataset will be used for secondary observational studies such as this one, addressing additional hypothesis-driven questions (e.g., risk factors for gastrointestinal bleeding). Access by REVISE investigators will follow a submitted rationale, analysis plan and approval by the Management Committee. Requests for access to the dataset by external investigators will be considered following a submitted rationale, analysis plan and approval by the Management Committee and research ethics boards as relevant. Requirements will be stipulated in a pre-specified data sharing agreement. Only de-identified data will be provided and will be transferred via a secure web portal.

